# The immediate psychological response of the general population in Saudi Arabia during COVID-19 pandemic: a cross-sectional study

**DOI:** 10.1101/2020.06.19.20135533

**Authors:** Royes Joseph, Dhfer Alshayban, Jisha M Lucca, Yasir Abdulaziz Alshehry

## Abstract

**Background:** The health and economic burden of pandemic diseases significantly cause psychosocial problems. The outbreak of COVID-19 may differentially exacerbate anxiety and stress in people subjected to the real or perceived threat of the virus.

**Method:** An online cross-sectional survey was carried out to assess the general population’s immediate psychological response during the initial state of the outbreak in Saudi Arabia. The study used brief screening tools PHQ-4 for anxiety-depression symptoms and IES-6 for posttraumatic stress disorder symptoms.

**Results:** Among the 584 respondents, 19.8% and 22.0% reported moderate to severe anxiety and depression symptoms respectively. According to the combined PHQ-4 score, 14.5% of participants showed moderate to severe anxiety or depression disorder. Overall, 64.8% met the level of clinical concern for posttraumatic stress disorder and 51.3% met the level of probable posttraumatic stress disorder diagnosis. Multivariate analyses showed that females, non-Saudi nationalities, and those who had a history of mental illness were more vulnerable to anxiety and depression disorders than their counterparts, whereas a higher prevalence of distress symptoms was reported among those who prefer Arabic over English for communication. It was found that people whose colleagues or family infected with the disease were more likely to report moderate to severe symptoms of anxiety or depression and distress. The study further showed that the higher the perceived threat, the higher the chances of exhibiting anxiety-depressive disorder symptoms and distress symptoms.

**Conclusion:** The findings might be a matter for serious concern, and considerable attention is required from authorities and policymakers regarding early detection and treatment of these illnesses in order to reduce the burden of the pandemic related mental illness.

## Introduction

Severe acute respiratory syndrome coronavirus 2 (SARS-CoV-2), publicly known as Coronavirus disease 2019 (COVID-19) or the novel coronavirus (2019-nCoV) disease, started as a zoonotic transmission event in China. Quickly it becomes a threat to the world community, eventually WHO declared COVID-19 as a pandemic disorder (World Health Organization, 2020a; World Health Organization, 2020b). Despite the extreme preventive measures, the number of people affected directly or indirectly is substantial, with no exemption to countries in the Arab region. As of 31^st^ May 2020, more than half a million COVID-19 cases, including 12353 deaths, were reported in the Arab region. The Kingdom of Saudi Arabia (KSA) was placed on the top after Iran in terms of the total number of COVID-19 confirmed cases (83384 cases) and on the bottom four with the lowest case fatality ratio of 0.60 (World Health Organization, 2020c).

Globally, governments and health services authorities are vigilant about the rapid spread, and they implement many measures to control the spread of SARS-CoV-2. These measures include identification and isolation of suspected and diagnosed cases, contact tracing and monitoring, the establishment of isolation units, practicing social distancing, home quarantine, travel restrictions, and the extreme stay-at-home restrictions (World Health Organization, 2020d). In KSA, a 16-days lockdown with travel restrictions was initially announced on 15^th^ Mar 2020 following the first COVID-19 case was reported on 2^nd^ Mar 2020, and it further extended for an indefinite period (Alshammari et al., 2020). Besides, a strict curfew with stay-at-home restriction was employed during April and May as the number of confirmed cases reached 1720 as of 1^st^ Apr and 240971 as of 1^st^ May (Alshammari et al., 2020). The steep increases in the number of confirmed cases and the restrictions on movement and day-to-day activities may negatively affect people’s mental health (World Health Organization, 2020e; World Health Organization, 2020f).

Previous studies on SARS and Ebola outbreaks reported a wide array of psychiatric morbidities, including anxiety, psychomotor excitement, panic attack, posttraumatic stress disorder (PTSD), psychotic symptoms, persistent depression, delirium, and even suicidality (Mak et al., 2010; Jalloh et al., 2018; Cabello et al., 2020; Rogers et al., 2020). Frontline health care professionals, especially those working in hospitals caring for people, high-risk communities, and the survivors were more vulnerable to the outbreak-related mental health problems. The largest outbreak of Middle East respiratory syndrome coronavirus (MERS-CoV) was seen in KSA with 2102 confirmed cases during the period 2012-2019 (World Health Organization, 2019). A previous study on MERS-CoV reported that residents in the country, especially health care providers, experienced a wide range of mental health problems (Alnajjar et al., 2016).

Infectious disease outbreaks significantly cause psychosocial problems (Rogers et al., 2020; World Health Organization, 2020e; World Health Organization, 2020f). The outbreak of COVID-19 may differentially exacerbate anxiety and stress in people subjected to the real or perceived threat of the virus. The uncertainty of the incubation period, asymptomatic transmission, extraordinary large-scale quarantine measures, curfew, and lockdown surge the likelihood of adverse psychosocial effects on the public (World Health Organization, 2020d). Fear and anxiety about a disease can be overwhelming and cause strong emotions in adults and children. The extensive media coverage that highlights COVID-19 as a unique threat may further added to panic, stress, and the potential for hysteria. The WHO, along with national authorities, is keen on monitoring the impact of the COVID-19 pandemic on people’s mental health (World Health Organization, 2020e; World Health Organization, 2020f). Epidemiological documents on the psychiatric morbidity of the current COVID-19 pandemic are limited in the Arab region. In response to this call, this study was aimed to collect information on the mental health problem and psychiatric comorbidities during the pandemic period among the general population of the KSA. The objectives of this study were (1) to estimate the prevalence of anxiety-depression and distress following exposure to the COVID-19 outbreak in the general population; and (2) to assess the impact of COVID-19 experience and the perceived threat on anxiety, depression, and distress symptoms.

## Methodology

### Study design

An online-based cross-sectional study was carried out from 12^th^ April to 10^th^ May 2020. During the period, the number of confirmed COVID-19 cases was increased from 4462 to 39048, and the number of related deaths was increased from 59 to 246 (World Health Organization, 2020g). The online questionnaire was created using the QuestionPro tool and circulated through emails and social media platforms. For the data collection, the administrative regions of the KSA were merged into five geographical regions (Eastern, Central, Western, Northern, and Southern regions). The incidence of COVID-19 was higher in the Eastern, Central, and Western regions compared to the other regions. The distribution of the questionnaires was planned to have participants from these regions. Participation in the study was voluntary, and the participants were requested to provide their consent and continue with the questionnaire items by selecting a checkbox. Residents of Saudi Arabia who were aged 15 years or older were eligible to participate in the study if they could respond to the questionnaire either in Arabic or English. The study was approved by the Institutional Review Board (IRB approval number: IRB-2020-05-173).

### Sample size

As the study expected a high prevalence of mental health symptoms, a prevalence of 50% that yields the largest sample size was assumed. The study required sample size of 600 participants to detect the prevalence with a 95% confidence level and a 4% type 1 error rate.

### Data collection

Both Arabic and English version of the study questionnaire was made available through QuestionPro. The study adopted a questionnaire that assessed the mental health impact of the Ebola epidemic in the general population of Sierra Leone (Jalloh et al., 2018), with modifications suitable for COVID-19 in the Middle-East population. The questionnaire has 30 items over six sections. The sections include 1) socio-demographic and clinical characteristics of participants; 2) personal experience with the COVID-19 related events (knowing persons tested positive for SARS-CoV-2); 3) perceived COVID-19 threat to the country, neighborhood, and household; 4) anxiety and depression using the Patient Health Questionnaire 4 (PHQ-4) (Kroenke et al., 2009); and 5) distress using Impact of Event Scale – 6 (IES-6) (Thoresen et al., 2010; Giorgi et al., 2015).

PHQ-4 is a validated ultra-brief screening scale consisting of the first two items from the PHQ-9 questionnaire (a 9-item depression severity scale), and generalized anxiety disorder −7 (GAD-7; a 7-item generalized anxiety severity scale), respectively (Kroenke et al., 2009). Both Arabic and English versions of PHQ-4 were obtained from the PHQ website (https://www.phqscreeners.com/select-screener). Each subscale ranges from a score of 0-6, and the combined scale ranges from a score of 0-12. Previous research suggests scores of three or greater as moderate and five or greater as severe indicators for potential cases of depression and anxiety in the depression and anxiety subscales, respectively (Löwe et al., 2010). For use in clinical practice, the study suggested considering the combined scale with scores of six or greater as “yellow flags-moderate” and scores of nine or greater as “red flags - severe” for the presence of a depressive or an anxiety disorder (Löwe et al., 2010).

IES-6 is a brief scale measure severity of distress following exposure to traumatic events, and it has six items that were measured on a 4-point severity scale with a total score ranges from a scale of 0-24, which reflects the level of posttraumatic stress reactions (Thoresen et al., 2010; Giorgi et al., 2015). The scale is particularly useful in studies for quick screening of a large population for PTSD and was used in an Ebola epidemic study (Jalloh et al., 2018). We used scores of 7 or greater as a ‘clinical concern’ for PTSD and scores of 9 or greater as ‘probable diagnosis’ of PTSD according to a previous study (Jalloh et al., 2018). The Arabic instrument was validated through translation and back-translation method by three independent groups of translators. The Arabic version was piloted on a sample after consonance was achieved between translators. The reliability test gave a Cronbach’s α of 0.79.

### Analysis

Data were summarized using frequencies (percentages) and mean (standard deviation) as appropriate. For analyses, binary variables were created for indicating the presence of symptoms for anxiety-depression and clinical concerns for PTSD, respectively. Univariate analyses were carried out using the Chi-square test for testing the association between predictors and the outcome variables. Multivariate logistic regression models were used to assess the determinants of psychological disorder symptoms. Variables that found significant in the univariate analysis were entered into multivariable logistic regression. The adjusted odds ratio (AOR) and its 95% confidence interval (CI) were reported. A two-sided p-value of 0.05 or lower regarded as statistically significant. All analyses were carried out using Stata/SE 16.1.

## Results

### Characteristics of participants

The questionnaire was reached to 1273 individuals, and 60% (n=765) of them initiated filling the questionnaire, and 649 met the eligibility criteria. A total of 90% (584/649) completed the baseline and PHQ-4 items, and 50 of those completed the PHQ-4 items had discontinued before filling the IES-6 related items.

The participants’ characteristics are summarized in Table 1. Majority of participants were males, aged less than 45 years, Saudi nationalities, and mostly from the Eastern, Central or Western regions. Importantly, one-fifth of participants were health care professionals, and nearly one-third were working from the office site during the COVID-19 lockdown. A minor proportion of participants reported that they had a serious or chronic illness, or some mental health issues before the COVID-19 outbreak. Majority preferred the Arabic version of the questionnaire over the English version, and there observed an increased participation over weeks.

**Table 1:** Characteristics of participants (n=584)

| Baseline data | n | % |
| --- | --- | --- |
| Gender |  |  |
| Male | 361 | 61.8% |
| Female | 223 | 38.2% |
| Age |  |  |
| <25 years | 132 | 22.6% |
| 25-34 years | 232 | 39.7% |
| 35-44 years | 107 | 18.3% |
| 45 years or older | 113 | 19.3% |
| Nationality |  |  |
| Non-Saudi | 124 | 21.2% |
| Saudi | 460 | 78.8% |
| Region of Saudi |  |  |
| Eastern Region | 250 | 42.8% |
| Central Region | 91 | 15.6% |
| Western Region | 133 | 22.8% |
| Southern/northern Region | 110 | 18.8% |
| Education |  |  |
| Secondary school or lower | 126 | 21.6% |
| Bachelor's degree | 312 | 53.4% |
| Professional degree, masters or higher | 146 | 25.0% |
| Employment Status |  |  |
| Student | 112 | 19.2% |
| Health Professionals | 110 | 18.8% |
| Other government sectors | 154 | 26.4% |
| Other private sectors | 134 | 22.9% |
| Unemployed | 74 | 12.7% |
| Working status during the lockdown |  |  |
| Not working | 160 | 27.4% |
| Mostly work-from-home | 244 | 41.8% |
| Mostly work at the office site | 180 | 30.8% |
| Marital status |  |  |
| Single | 256 | 43.8% |
| Married | 328 | 56.2% |
| Accommodation status |  |  |
| Living alone | 57 | 9.8% |
| Living with family | 502 | 86.0% |
| Living in shared accommodation | 25 | 4.3% |
| Presence chronic illness |  |  |
| No | 518 | 88.7% |
| Yes | 66 | 11.3% |
| Existing mental health issues |  |  |
| No | 564 | 96.6% |
| Yes | 20 | 3.4% |
| Preferred communication language |  |  |
| Arabic | 423 | 72.4% |
| English | 161 | 27.6% |
| Week of data collection |  |  |
| Week 1 | 74 | 12.7% |
| Week 2 | 162 | 27.7% |
| Week 3 | 146 | 25.0% |
| Week 4 | 202 | 34.6% |

### Prevalence of Anxiety and depression

A total of 104 (17.6%) and 128 (21.9%) participants showed moderate-severe anxiety and depressive symptoms, respectively (Figure 1). According to the combined scale PHQ-4, prevalence of moderate-severe anxiety-depressive disorder symptom was 14.5% (n=85). Importantly, 20 (3.4%) were on the severe anxiety-depressive disorder as per the combined scale. The mean (SD) PHQ-4 score was 2.9 (2.6) on a 0-12 scale.

**Figure 1:**
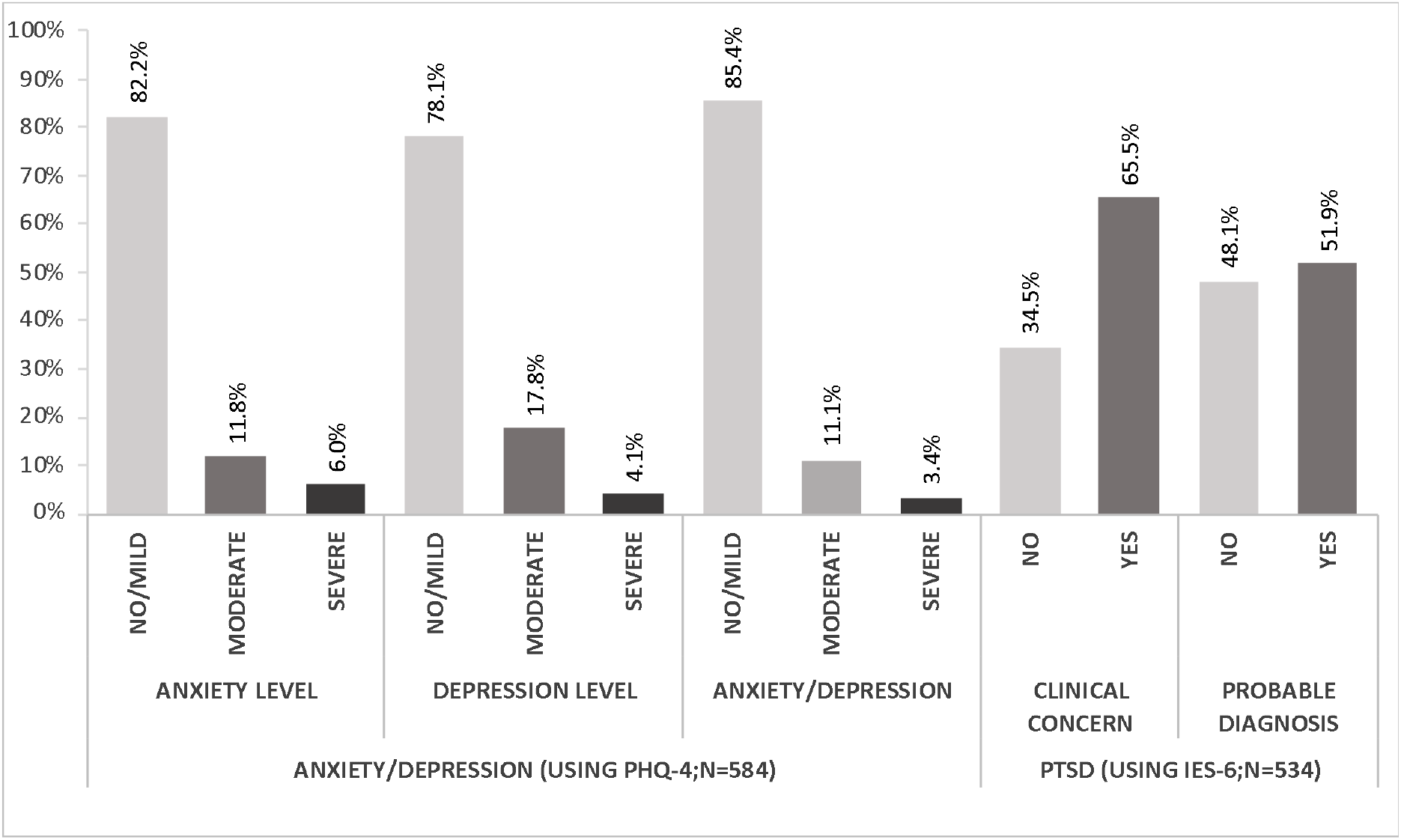
Prevalence of anxiety/depression and distress

Univariate analyses showed gender and existing mental health issues were significantly associated with the moderate-severe anxiety-depressive disorder (Columns 2, 3 & 4 in Table 2). A multivariate logistic regression model showed a higher risk of the moderate-severe anxiety-depressive disorder among females [AOR (95% CI): 2.8 (1.7, 4.6)] compared to males, non-Saudi individuals [AOR (95% CI): 1.8 (1.03, 3.2)] compared to Saudi nationals, and individuals having existing mental health issues [AOR (95% CI): 4.0 (1.4, 10.9)] compared to those without any existing mental health issues. The higher risk, but not statistically significant, was also observed among students [AOR (95% CI): 1.8 (0.75, 4.3)] and health professionals [AOR (95% CI): 2.1 (0.86, 4.9)] compared to unemployed.

**Table 2:**
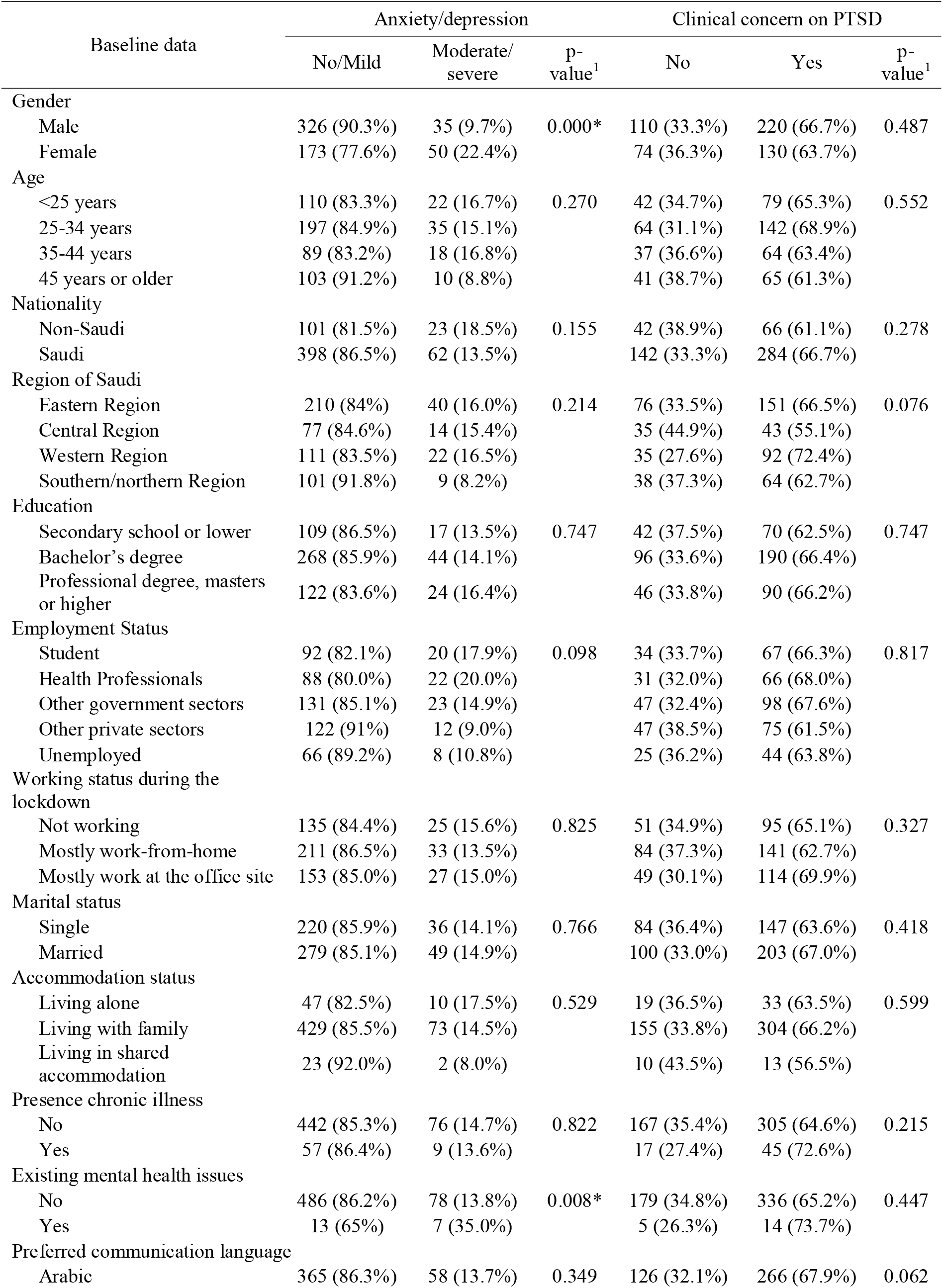
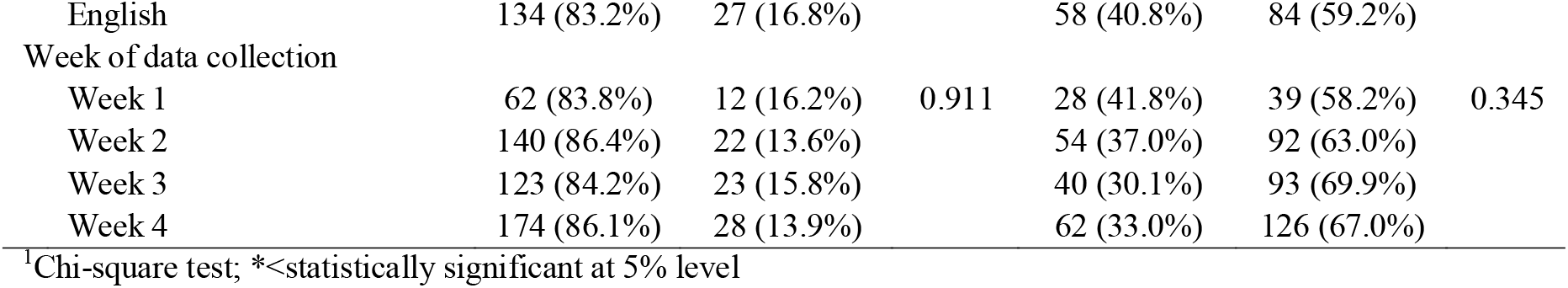
Univariate analysis – association with socio-demographic characteristics

| Baseline data | Anxiety/depression |  |  | Clinical concern on PTSD |  |  |
| --- | --- | --- | --- | --- | --- | --- |
|  | No/Mild | Moderate/<br>severe | p-value<sup>1</sup> | No | Yes | p-value<sup>1</sup> |
| Gender |  |  |  |  |  |  |
| Male | 326 (90.3%) | 35 (9.7%) | 0.000* | 110 (33.3%) | 220 (66.7%) | 0.487 |
| Female | 173 (77.6%) | 50 (22.4%) |  | 74 (36.3%) | 130 (63.7%) |  |
| Age |  |  |  |  |  |  |
| <25 years | 110 (83.3%) | 22 (16.7%) | 0.270 | 42 (34.7%) | 79 (65.3%) | 0.552 |
| 25-34 years | 197 (84.9%) | 35 (15.1%) |  | 64 (31.1%) | 142 (68.9%) |  |
| 35-44 years | 89 (83.2%) | 18 (16.8%) |  | 37 (36.6%) | 64 (63.4%) |  |
| 45 years or older | 103 (91.2%) | 10 (8.8%) |  | 41 (38.7%) | 65 (61.3%) |  |
| Nationality |  |  |  |  |  |  |
| Non-Saudi | 101 (81.5%) | 23 (18.5%) | 0.155 | 42 (38.9%) | 66 (61.1%) | 0.278 |
| Saudi | 398 (86.5%) | 62 (13.5%) |  | 142 (33.3%) | 284 (66.7%) |  |
| Region of Saudi |  |  |  |  |  |  |
| Eastern Region | 210 (84%) | 40 (16.0%) | 0.214 | 76 (33.5%) | 151 (66.5%) | 0.076 |
| Central Region | 77 (84.6%) | 14 (15.4%) |  | 35 (44.9%) | 43 (55.1%) |  |
| Western Region | 111 (83.5%) | 22 (16.5%) |  | 35 (27.6%) | 92 (72.4%) |  |
| Southern/northern Region | 101 (91.8%) | 9 (8.2%) |  | 38 (37.3%) | 64 (62.7%) |  |
| Education |  |  |  |  |  |  |
| Secondary school or lower | 109 (86.5%) | 17 (13.5%) | 0.747 | 42 (37.5%) | 70 (62.5%) | 0.747 |
| Bachelor's degree | 268 (85.9%) | 44 (14.1%) |  | 96 (33.6%) | 190 (66.4%) |  |
| Professional degree, masters or higher | 122 (83.6%) | 24 (16.4%) |  | 46 (33.8%) | 90 (66.2%) |  |
| Employment Status |  |  |  |  |  |  |
| Student | 92 (82.1%) | 20 (17.9%) | 0.098 | 34 (33.7%) | 67 (66.3%) | 0.817 |
| Health Professionals | 88 (80.0%) | 22 (20.0%) |  | 31 (32.0%) | 66 (68.0%) |  |
| Other government sectors | 131 (85.1%) | 23 (14.9%) |  | 47 (32.4%) | 98 (67.6%) |  |
| Other private sectors | 122 (91%) | 12 (9.0%) |  | 47 (38.5%) | 75 (61.5%) |  |
| Unemployed | 66 (89.2%) | 8 (10.8%) |  | 25 (36.2%) | 44 (63.8%) |  |
| Working status during the lockdown |  |  |  |  |  |  |
| Not working | 135 (84.4%) | 25 (15.6%) | 0.825 | 51 (34.9%) | 95 (65.1%) | 0.327 |
| Mostly work-from-home | 211 (86.5%) | 33 (13.5%) |  | 84 (37.3%) | 141 (62.7%) |  |
| Mostly work at the office site | 153 (85.0%) | 27 (15.0%) |  | 49 (30.1%) | 114 (69.9%) |  |
| Marital status |  |  |  |  |  |  |
| Single | 220 (85.9%) | 36 (14.1%) | 0.766 | 84 (36.4%) | 147 (63.6%) | 0.418 |
| Married | 279 (85.1%) | 49 (14.9%) |  | 100 (33.0%) | 203 (67.0%) |  |
| Accommodation status |  |  |  |  |  |  |
| Living alone | 47 (82.5%) | 10 (17.5%) | 0.529 | 19 (36.5%) | 33 (63.5%) | 0.599 |
| Living with family | 429 (85.5%) | 73 (14.5%) |  | 155 (33.8%) | 304 (66.2%) |  |
| Living in shared accommodation | 23 (92.0%) | 2 (8.0%) |  | 10 (43.5%) | 13 (56.5%) |  |
| Presence chronic illness |  |  |  |  |  |  |
| No | 442 (85.3%) | 76 (14.7%) | 0.822 | 167 (35.4%) | 305 (64.6%) | 0.215 |
| Yes | 57 (86.4%) | 9 (13.6%) |  | 17 (27.4%) | 45 (72.6%) |  |
| Existing mental health issues |  |  |  |  |  |  |
| No | 486 (86.2%) | 78 (13.8%) | 0.008* | 179 (34.8%) | 336 (65.2%) | 0.447 |
| Yes | 13 (65%) | 7 (35.0%) |  | 5 (26.3%) | 14 (73.7%) |  |
| Preferred communication language |  |  |  |  |  |  |
| Arabic | 365 (86.3%) | 58 (13.7%) | 0.349 | 126 (32.1%) | 266 (67.9%) | 0.062 |
| English | 134 (83.2%) | 27 (16.8%) |  | 58 (40.8%) | 84 (59.2%) |  |
| Week of data collection |  |  |  |  |  |  |
| Week 1 | 62 (83.8%) | 12 (16.2%) | 0.911 | 28 (41.8%) | 39 (58.2%) | 0.345 |
| Week 2 | 140 (86.4%) | 22 (13.6%) |  | 54 (37.0%) | 92 (63.0%) |  |
| Week 3 | 123 (84.2%) | 23 (15.8%) |  | 40 (30.1%) | 93 (69.9%) |  |
| Week 4 | 174 (86.1%) | 28 (13.9%) |  | 62 (33.0%) | 126 (67.0%) |  |

<sup>1</sup>Chi-square test; \*<statistically significant at 5% level

### Prevalence of posttraumatic stress disorder

The mean (SD) IES-6 score was 8.8 (4.9) on a 0-24 scale. The results in Figure 1 displays the prevalence of distress. Overall, 65.5% (n=350) met levels of clinical concern for PTSD and 51.9% (n=277) met levels of probable PTSD diagnosis.

Univariate analysis (Columns 5, 6 & 7 in Table 2) showed the risk of having clinical concern for PTSD was not statistically differed by the levels of socio-demographic characteristics of individuals. Multivariate analysis showed that only the variable preferred language for communication was found to be associated with the clinical concern for PTSD: more people who prefer the Arabic language at the risk compared to those prefer the English language [AOR (95% CI): 1.5 (1.01, 2.3)].

### Impact of personal experience with the COVID-19 related events on psychological disorders

As shown in Table 3, more than a quarter of the participants knew a person who was tested positive for SARS-CoV-2. Both univariate and multivariate analyses (model 1) showed that individuals having personal experience with COVID-19 related events were at higher risk for anxiety-depression [AOR (95% CI): 1.6 (0.94, 2.7)] and PTSD [AOR (95% CI): 1.7 (1.1, 2.6)]. Model 2 in Table 3 explored the association between the degree of personal level experience with COVID-19 and psychological disorders. The results showed that individuals who knew only non-hospitalized COVID-19 cases were at higher risk for anxiety-depression [prevalence=21.2%; AOR (95% CI): 2.0 (1.1, 3.5)] and individuals who knew hospitalized COVID cases were at higher risk for PTSD [prevalence=80.4%; AOR (95% CI): 2.5 (1.2, 5.0)] compared to who did not have such experience levels (prevalence=12.8% for anxiety-depression and 62.6% for PTSD symptoms).

**Table 3:**
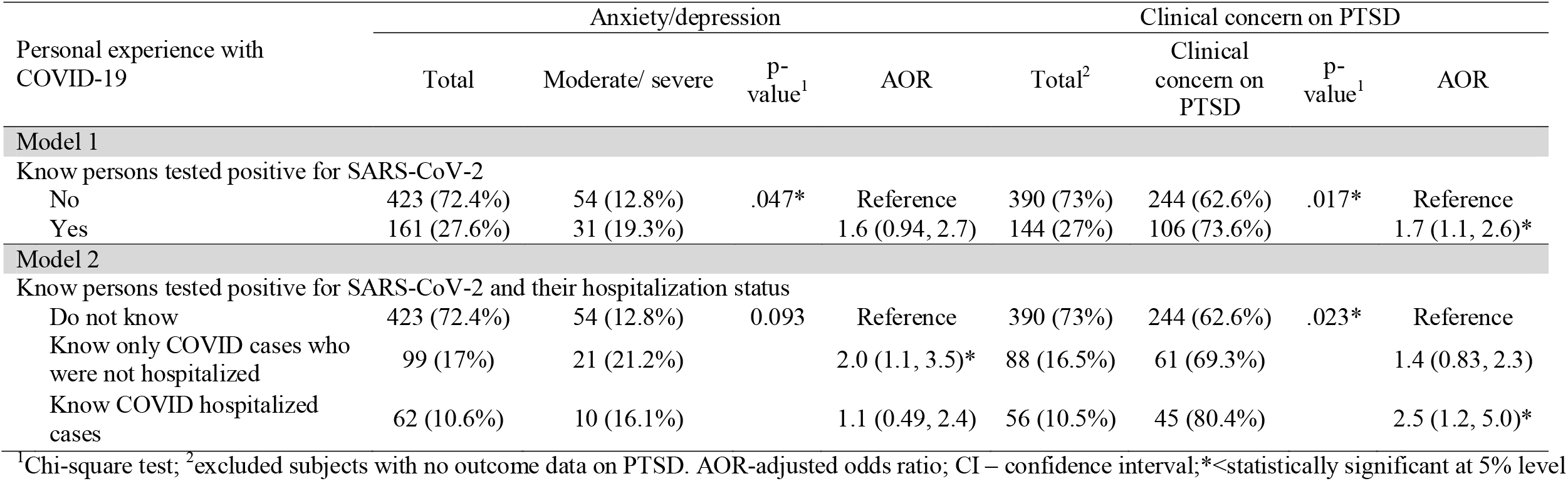
Personal experience with the COVID-19 related events

| Personal experience with COVID-19 | Anxiety/depression |  |  |  | Clinical concern on PTSD |  |  |  |
| --- | --- | --- | --- | --- | --- | --- | --- | --- |
|  | Total | Moderate/ severe | p-value<sup>1</sup> | AOR | Total<sup>2</sup> | Clinical concern on PTSD | p-value<sup>1</sup> | AOR |
| **Model 1** |  |  |  |  |  |  |  |  |
| Know persons tested positive for SARS-CoV-2 |  |  |  |  |  |  |  |  |
| No | 423 (72.4%) | 54 (12.8%) | .047* | Reference | 390 (73%) | 244 (62.6%) | .017* | Reference |
| Yes | 161 (27.6%) | 31 (19.3%) |  | 1.6 (0.94, 2.7) | 144 (27%) | 106 (73.6%) |  | 1.7 (1.1, 2.6)* |
| **Model 2** |  |  |  |  |  |  |  |  |
| Know persons tested positive for SARS-CoV-2 and their hospitalization status |  |  |  |  |  |  |  |  |
| Do not know | 423 (72.4%) | 54 (12.8%) | 0.093 | Reference | 390 (73%) | 244 (62.6%) | .023* | Reference |
| Know only COVID cases who were not hospitalized | 99 (17%) | 21 (21.2%) |  | 2.0 (1.1, 3.5)* | 88 (16.5%) | 61 (69.3%) |  | 1.4 (0.83, 2.3) |
| Know COVID hospitalized cases | 62 (10.6%) | 10 (16.1%) |  | 1.1 (0.49, 2.4) | 56 (10.5%) | 45 (80.4%) |  | 2.5 (1.2, 5.0)* |

<sup>1</sup>Chi-square test; <sup>2</sup>excluded subjects with no outcome data on PTSD. AOR-adjusted odds ratio; CI – confidence interval; \*<statistically significant at 5% level

### Impact of perceived COVID-19 threat on psychological disorders

Table 4 lists the prevalence of moderate-severe anxiety-depression and PTSD by different levels of perceived COVID-19 threat. Model 1 explored the association of the perceived threat at the level of the country, neighborhood and household with the psychological disorders. The results showed that the moderate-severe perceived threat at the household level was significantly associated with anxiety-depressive symptoms [AOR (95% CI): 2.3 (1.01, 5.1)] and PTSD symptoms [AOR (95% CI): 2.4 (1.4, 4.1)]. Model 2 considered a composite risk perception score as a predictor for the psychological disorder symptoms. According to the model 2, persons with moderate-severe threat perception at all levels were at a substantially higher risk of reporting moderate-severe anxiety-depression symptoms [prevalence=20.5%; AOR (95% CI): 9.0 (2.1, 38.8)] and PTSD symptoms [prevalence=74.5%; AOR (95% CI): 4.5 (2.4, 8.1)] compared to those with minor or lower threat perception at all levels (2.8% for anxiety-depression and 40.0% for PTSD symptoms).

**Table 4:** Perceived COVID-19 Threat

| Perceived COVID-19 Threat | Anxiety/depression |  |  |  | Clinical concern on PTSD |  |  |  |
| --- | --- | --- | --- | --- | --- | --- | --- | --- |
|  | Total | Moderate/ severe | p-value<sup>1</sup> | AOR | Total<sup>2</sup> | Clinical concern on PTSD | p-value<sup>1</sup> | AOR |
| **Model 1** |  |  |  |  |  |  |  |  |
| COVID-19 is a threat for the country |  |  |  |  |  |  |  |  |
| Not a threat | 39 (6.7%) | 3 (7.7%) | .047* | Reference | 32 (6%) | 13 (40.6%) | .001* | Reference |
| Minor | 55 (9.4%) | 3 (5.5%) |  | 0.7 (0.1, 4.6) | 48 (9%) | 26 (54.2%) |  | 1.4 (0.5, 3.9) |
| Moderate/Severe | 490 (83.9%) | 79 (16.1%) |  | 1.3 (0.3, 6.3) | 454 (85%) | 311 (68.5%) |  | 1.5 (0.6, 3.9) |
| COVID-19 is a threat in your neighborhood |  |  |  |  |  |  |  |  |
| Not a threat | 43 (7.4%) | 3 (7%) | .025* | Reference | 37 (6.9%) | 13 (35.1%) | .000* | Reference |
| Minor | 76 (13%) | 5 (6.6%) |  | 0.9 (0.2, 5.4) | 68 (12.7%) | 36 (52.9%) |  | 1.4 (0.5, 3.7) |
| Moderate/Severe | 465 (79.6%) | 77 (16.6%) |  | 1.1 (0.2, 6.1) | 429 (80.3%) | 301 (70.2%) |  | 1.7 (0.7, 4.6) |
| COVID-19 is a threat in your family |  |  |  |  |  |  |  |  |
| Not a threat | 150 (25.7%) | 12 (8%) | .000* | Reference | 135 (25.3%) | 67 (49.6%) | .000* | Reference |
| Minor | 137 (23.5%) | 12 (8.8%) |  | 0.9 (0.4, 2.4) | 127 (23.8%) | 80 (63%) |  | 1.5 (0.9, 2.6) |
| Moderate/Severe | 297 (50.9%) | 61 (20.5%) |  | 2.3 (1.01, 5.1)* | 272 (50.9%) | 203 (74.6%) |  | 2.4 (1.4, 4.1)* |
| Model 2 |  |  |  |  |  |  |  |  |
| Risk perception - Composite |  |  |  |  |  |  |  |  |
| minor/no threat at all levels | 71 (12.2%) | 2 (2.8%) | .000* | Reference | 60 (11.2%) | 24 (40.0%) | .000* | Reference |
| severe threat at some level, but not at all levels | 230 (39.4%) | 25 (10.9%) |  | 4.4 (1.01, 19.7)* | 215 (40.3%) | 133 (61.9%) |  | 2.4 (1.3, 4.3)* |
| severe threat at all levels | 283 (48.5%) | 58 (20.5%) |  | 9.0 (2.1, 38.8)* | 259 (48.5%) | 193 (74.5%) |  | 4.5 (2.4, 8.1)* |

<sup>1</sup>Chi-square test; <sup>2</sup>excluded subjects with no outcome data on PTSD. AOR-adjusted odds ratio; CI – confidence interval; \*<statistically significant at 5% level

## Discussion

The study assessed the prevalence of psychological symptoms using the brief screening tools PHQ-4 for anxiety-depression symptoms (Kroenke et al., 2009) and IES-6 for distress symptoms (Thoresen et al., 2010; Giorgi et al., 2015) among the general population in KSA during the initial period of COVID-19. In this study, 19.8% and 22.0% of respondents reported moderate to severe anxiety and depression symptoms, respectively. According to the combined PHQ-4 score, which was suggested for the screening purpose (Löwe et al., 2010), 14.5% of participants showed symptoms of moderate to severe anxiety or depression disorder. Overall, 64.8% of participants met the level of clinical concern for PTSD, and 51.3% met the level of probable PTSD diagnosis. The results indicate that the COVID-19 pandemic may cause significant psychiatric burdens among the general population during the outbreak, as evident from the previous coronavirus epidemics (Rogers et al., 2020).

The prevalence of anxiety and depression disorders among the general public was higher than that reported by a study conducted during the same period in the Kingdom (Alyami et al., 2020). However, the prevalence was within the range of that reported by studies among the various population within Saudi Arabia (prevalence varied 15.8% to 26% for depression and 16.6% to 66% for anxiety) before the COVID-19 outbreak (Al-Qadhi et al., 2014; Alzahrani et al., 2016; Alanazy, 2019; Alharithy et al., 2019; Al Salman et al., 2020). A similar or higher prevalence of psychological burden was reported during the past major infectious diseases outbreaks, such as MERS, SARS and Ebola, especially among those directly affected by the outbreaks (Mak et al., 2010; Alnajjar et al., 2016; Jalloh et al., 2018; Cabello et al., 2020; Brooks et al., 2020). A recent systematic review showed that nearly one-third of patients admitted to hospital due to SARS or MERS exhibited anxiety and depression symptoms during the acute illness stage, but the prevalence reduced to 15% during the post-illness stage (Rogers et al., 2020). Similarly, studies from China reported that a significant minority of the adult population exhibited severe-moderate anxiety and depression symptoms during the COVID-19 pandemic (Wang et al., 2020; Zhao et al., 2020).

Researches on previous infectious disease outbreak warn about psychiatric presentations, including posttraumatic stress reactions, associated with the COVID-19 pandemic (Jalloh et al., 2018; Rogers et al., 2020; Brooks et al., 2020; Xiao et al., 2020). The estimate of the prevalence of distress from the present study indicated that more than half of the general public in Saudi Arabia were under severe distress during the pandemic period. The estimated prevalence among the general public was greater than what observed among workers attending emergency medical and fire services in the country (Qumri and Osman, 2014; Alghamdi et al., 2017; Alaqeel et al., 2019). A study from China also reported such a high prevalence among the general public during the current pandemic (Wang et al., 2020). This finding is a matter for serious concern as psychological problems such as PTSD need early detection and intervention. The high prevalence may be the sudden reaction to the ongoing stressful time and the continuous insensitivity around it due to the most severe pandemic ever had happened in the last decade. The nationwide lockdown and associated social and financial uncertainty, poor understanding of the virus and spreading mechanisms, uncertainty about the vaccination and treatment, and fear of getting infected and transmitting the virus to family members or friends also may stimulate the distress.

It is well reported that natural disasters are typically unpredictable and leaves the victims in a state of shock (Sajid, 2007). Currently, the viral pandemic is spookily similar to a natural disaster or making things worse as no vaccine has been developed, or there are no specific treatments. Similarly, the public has been urged to practice challenging preventative measures, particularly self-isolation and social distancing (Nasrallah, 2020). Through various media during the pandemic period, people are continuously exposed to massive information on COVID-19 related events in the country and the world. Importantly, the public sense massive hoax information from social media and channeled fear into frantic buying and hoarding of food and non-food items (Nasrallah, 2020). Individuals experiencing these sudden upheavals will undergo biological and psychological effects and activate the body’s “fight or flight or freeze” response (Schmidt et al., 2008). If the flight or fight response activated continuously and definitely, that might result in anxiety (Retano, 2014). It has also been reported that sustained or chronic tread can eventually lead to depression. Similarly, persons who are in acute stress disorder are likely to display subsequent PTSD (Maeng and Milad, 2017). The much higher prevalence of distress may be due to the IES-6 items being more specific to the COVID-19 pandemic, while the PHQ-4 is not.

Multivariate analysis of our data showed that females, non-Saudi nationalities, and those with mental illness histories are more vulnerable to anxiety and depression disorders than their counterparts. The prevalence of the mental disorder symptoms was also found to be higher, but not significantly, among students and health professionals. Previous epidemiological studies on endemics have demonstrated that women are vulnerable to anxiety and depression (Rogers et al., 2020). Recent reports from the Ministry of Health revealed that more than three-quarters of confirmed cases of COVID-19 were from immigrant workers. The higher COVID-19 incidence among expatriates may reflect on their mental health as well. Factors such as living in a crowded area, public transportation to work, fear of losing a job, consequent deprivation of their income, unpredictable future during and after the current pandemic, and the pandemic situation in their native country may explain the higher prevalence of mental health issues among the non-Saudi nationalities. People with existing mental health conditions could be more substantially predisposed by the emotional responses brought on by the COVID-19 epidemic, resulting in relapses or worsening of an already existing mental health condition (Yao et al., 2020). Similar to the results reported by a study from China (Wang et al., 2020), students were found to experience a high level of anxiety or depression. In Saudi Arabia, the pandemic outbreak was in the middle of the second semester. The government closed all the educational facilities and switched them to remote learning. Thus, the students faced some obstacles, such as new methods of teaching, different exam-style, and lack of appropriate devices for learning. Also, uncertainty about academic progression and fear of losing the year or occurrence of delays in their studies could negatively impact students’ mental health. Similarly, our data showed that one in four healthcare professionals had anxiety or depression symptoms. A recent meta-analysis demonstrated mental health illness are more prevalent among health care workers than in general public during or after infectious disease outbreaks (Cabello et al., 2020). In terms of PTSD in the present study, the immediate psychological response was similarly prevalent across all considered socio-demographic factors except the preferred language for communication. Since most of the COVID-19 related communications are in Arabic, people prefer Arabic may be more exposed to COVID-19 related events. It could be a reason that led to a higher prevalence of distress symptoms among those who prefer Arabic over English for communication.

The present study also explored how individuals’ personal experience with pandemic-related events affects their mental health during the outbreak. It was found that people whose colleagues or family infected with SARS-CoV-2 were more likely to report moderate to severe symptoms of anxiety, depression, and PTSD. Further, it was observed that knowing COVID-19 cases had more than double the chance for showing symptoms of PTSD. A similar finding was observed in the past as well as in the current outbreaks (Jalloh et al., 2018; Wang et al., 2020). The higher prevalence could be due to concern over the well-being of family and colleagues, and the fear of personal safety.

More than three-fourth of participants thought COVID-19 is a moderate-severe threat to their country and neighborhood, while nearly 50% believed the pandemic is a minor or no threat to their family. The study showed that the higher the perceived threat indicate higher the chances for exhibiting anxiety-depressive disorder symptoms and distress symptoms. The finding was more evident on the household level perceived threat data compared to the threat at other levels. Further, a substantially higher proportion of participants who had perceived moderate-severe threat at all levels reported anxiety-depressive disorder symptoms (20.5% vs. 2.8%; AOR=9.0) and distress symptoms (74.% vs. 40.0%; AOR=4.5) compared to that among those who had perceived minor or no threat at all levels. Similar findings reported by an epidemiological study from china (Wang et al., 2020), wherein high levels of concern about other family members getting COVID-19 were significantly associated with higher stress scale scores. From the lessons learned from the MERS-CoV epidemic, Saudi Arabia started its public awareness program much before the first COVID-19 case reported on 2nd Mar, 2020, to prepare the people to cope with the pandemic and related mental health problems (Alshammari et al., 2020). The campaign employs television advertisements and different social media platforms. In addition to daily press releases, residents in the country receive daily text messages on preventive measures, the new developments of COVID-19, and the sources of help to deal with the pandemic related events (Alshammari et al., 2020). The accurate updates from authorities help to reduce the impact of rumors, still these measures could have adverse psychological effects during the early stage of the outbreak, where growth on the number of cases was significant. Though the prevalence of initial psychological responses may get reduce over time as the number of recovered cases increase, still a significant minority may need proper medical attention for a prolonged time as evident from past infectious disease outbreaks (Rogers et al., 2020; Xiao et al., 2020).

This study has several limitations. Firstly, the study had a limited sample size. Though the Kingdom of Saudi Arabia comprises 13 administrative regions but classified these regions into five geographical regions due to limited sample size, especially from the Northern or Southern regions. During the initial stage, the disease outbreak was substantial in the Western, Eastern, and Central regions, but not in the Northern or Southern regions. Secondly, the screening was done using self-administered questionnaires through an online tool, but on two languages. Therefore, the responders may provide data that meet the social expectation rather than reality, and the clinical significance may be unpredictable. Another potential limitation may be the oversampling of a particular network of similar groups, which may lead to selection bias. Further, the study did not exploit the psychological responses of COVID-19 patients or survivors as such as the primary focus of the study was the general public. Finally, but importantly, the study used ultra-brief questionnaires for the screening for anxiety, depression, and distress. However, the questionnaires are proven to be used for the initial screening of a large number of individuals (Kroenke et al., 2009; Thoresen et al., 2010; Löwe et al., 2010; Giorgi et al., 2015).

## Conclusion

The study provided a picture about the occurrence of depression, anxiety, and distress among the general public during the initial phase of the COVID-19 pandemic in Saudi Arabia. The study found that a minority but a substantial proportion of individuals had exhibited symptoms for anxiety or depression, while a majority reported symptoms for psychological distress. Personal experience with the disease-related events and perceived threat to the society and family were found associated with the symptoms of mental health illness. Though the severity of the pandemic related mental health illness may reduce over time as evident from the infectious disease outbreaks, considerable attention is required from authorities and policymakers regarding early detection and treatment of these illnesses in order to enhance the reduction further.

## Data Availability

The data that support the findings of this study are available from the corresponding author upon reasonable request.

## Contributors

RJ had the idea for the study. RJ, DA and JL equally contributed in study design and literature review. All authors had involved in the data collection. RJ did the data analysis. All authors interpreted the data analysis and helped in drafting the manuscript.

